# Immunodepletion with Hypoxemia: A Potential High Risk Subtype of Coronavirus Disease 2019

**DOI:** 10.1101/2020.03.03.20030650

**Authors:** Lilei Yu, Yongqing Tong, Gaigai Shen, Aisi Fu, Yanqiu Lai, Xiaoya Zhou, Yuan Yuan, Yuhong Wang, Yuchen Pan, Zhiyao Yu, Yan Li, Tiangang Liu, Hong Jiang

## Abstract

**Background:** The outbreak of COVID-2019 is becoming a global public health emergency. Although its basic clinical features have been reported, the dynamic characteristics of immune system in COVID-2019 patients, especially those critical patients with refractory hypoxemia, are not yet well understood. We aim to describe the dynamic characteristics of immune system in 3 critical patients with refractory hypoxemia, and discuss the relationship between hypoxemia severity and immune cell levels, and the changes of gut microbes of COVID-2019 patient.

**Methods:** This is a retrospective study from 3 patients with 2019-nCoV infection admitted to Renmin Hospital of Wuhan University, a COVID-2019 designated hospital in Wuhan, from January 31 to February 6, 2020. All patients were diagnosed and classified based on the Diagnosis and Treatment of New Coronavirus Pneumonia (6th edition) published by the National Health Commission of China^4^. We recorded the epidemiological history, demographic features, clinical characteristics, symptoms and signs, treatment and clinical outcome in detail. Blood samples were collected and we determined the expression levels of immune cells (CD3^+^ T cells, CD4^+^ T cells, CD8^+^ T cells, CD19^+^ B cells, and CD16^+^56^+^ NK cells) in different time points. Nanopore Targeted Sequencing was used to determine the alterations of gut microbiota homeostasis.

**Results:** Apart from the clinical features described previously^4^, we found that four patients had decreased immune cells and refractory hypoxemia during the hospitalization, and the severity of hypoxemia was strongly correlated to the expression levels of immune cells. Additionally, we found that the proportion of probiotics was significantly reduced, such as Bifidobacterium, Lactobacillus, and Eubacterium, and the proportion of conditioned pathogenic bacteria was significantly increased, such as Corynebacterium of Actinobacteria and Ruthenibacterium of Firmicutes. Notably, all patients died.

**Conclusions:** We discussed the dynamic characteristics of host immune system and the imbalance of gut microbiota in 3 critical patients with COVID-2019. Hypoxemia severity was closely related with host immune cell levels, and the vicious circle between immune disorder and gut microbiota imbalance may be a high risk of fatal pneumonia. To the best of our knowledge, this is the first study which revealing that immunodepletion with refractory hypoxemia is a potential high risk subtype of COVID-2019 and the vicious circle between immune disorder and gut dysbiosis may be a high risk of fatal pneumonia.

## Introduction

As of Feb 29, 2020, more than 80000 cases have been confirmed in China and other countries, including South Korea, Japan, Italy, and Iran. This global public health emergency is attracting most concern throughout the world.

Host immune response is essential for the clearance of coronavirus infection, and the balance between coronavirus and host immunity is key to viral pathogenesis and will ultimately determine infection outcome^1^. Coronaviruses encode numerous viral gene products to evade and suppress host immune response, such as SARS-associated coronavirus nonstructural protein 1, which is able to inhibit host interferon-dependent antiviral signaling^2^, as well as another SARS coronavirus protein named open reading frame-9b, which can suppress mitochondria of immune cells^3^. Coronavirus suppressing host immunity has been demonstrated, however, the alterations of immune system in patients with COVID-2019 are not yet well understood.

In this study, we aim to describe the dynamic characteristics of immune system in 3 critical patients with refractory hypoxemia, and discuss the relationship between hypoxemia severity and immune cell levels. We hope to reveal that immuno-depletion carries a higher risk of death for COVID-2019 patients.

## Methods

### Patients

This is a retrospective study from 3 patients with 2019-nCoV infection admitted to Renmin Hospital of Wuhan University, Wuhan, China, from January 31, 2020 to February 6, 2020. All patients were diagnosed and classified based on the Diagnosis and Treatment of New Coronavirus Pneumonia (7th edition) published by the National Health Commission of China^4^. We recorded the epidemiological history, demographic features, clinical characteristics, symptoms and signs, treatment and clinical outcome in detail. This study was reviewed and approved by the Medical Ethical Committee of Renmin Hospital of Wuhan University (approval number WDRY2020-K079).

### Laboratory characteristics

Nasopharyngeal-swab specimens were collected and maintained in viral-transport medium. 2019-nCoV was confirmed by real-time fluorescent RT-PCR. Positive confirmatory COVID-2019 patients were defined as those with both positive tests of 2019n-CoV open reading frame 1ab (2019n-CoV ORF1ab) and 2019n-Cov nucleocapsid protein gene (2019n-Cov N gene). Blood samples were collected from three patients at different time to determine the blood routine, coagulation function, blood biochemistry and inflammation-related markers. ECG monitors were used to record daily oximetry saturation of three patients. The lymphocyte test kit (Beckman Coulter Inc., FL, USA) were used to analyze the lymphocyte subsets including CD3^+^ T cells, CD4^+^ T cells, CD8^+^ T cells, CD19^+^ B cells, and also CD16^+^56^+^ NK cells in the periphery blood.

### Method of Nanopore Targeted Sequencing

The methods for these procedures are conducted as described in the Supplemental Appendix.

### Statistics

Categorical variables are expressed in percentage. Continuous variables were directly expressed as a range. To compare the differences in the gut microbiota between patients with health cohort which data were imported from Arumugam M’s^5^ research Comparative analysis was conducted by two tailed T test, Data analyses were performed using SPSS 23.0 software. P values < 0.05 were considered significant.

## Results

### Clinical presentation and laboratory findings

Three male adult patients diagnosed with COVID-2019 pneumonia were admitted to Renmin Hospital of Wuhan University on January 31, February 1 and 6, 2020 (Patient 1, Patient 2, Patient 3, respectively). The mean age of these patients are 74.3 ± 8.1 years. Patient 1 had a medical history of prostatic hyperplasia and Patient 3 had chronic bronchitis, while Patient 2 reported no underlying historic medical conditions. From the collection of clinical features, we found that all patients reported fever and dyspnea (Table 1). Moreover, Patient 1 also had diarrhea, and Patient 3 had cough and myalgia/fatigue additionally. Laboratory findings show that C-reactive protein (CRP) were obviously increased in all patients, and decreased lymphocyte counts and increased procalcitonin were observed in Patient 2 and 3 (Table 1). Additionally, Patient 2 also had elevated hypersensitive troponin I, creatine, and urea in serum, indicating myocardial injury and renal function damage. Patient 3 had increases in aspartate aminotransferase (ASL) and alanine aminotransferase (ALT) (Table 1), which indicate liver injury. During the hospitalization, all patients received empirical antiviral and antibiotic treatment (Table 1). Patient 1 also experienced intravenous immunoglobulin therapy from day 11 of hospitalization (Table 1). Notably, three patients died on day 24, 13, 12 of hospitalization, respectively.

**Table 1.** Clinical characteristics of 3 critical patients with COVID-2019.

|  |  |  |  |  |
| --- | --- | --- | --- | --- |
|  | **Patient1** | **Patient2** | **Patient3** | **n(%)** |
| **Basic individual information** |  |  |  |  |
| Date of admission | Feb 01 | Jan 31 | Feb 06 | .. |
| Age(years) | 65 | 80 | 78 | .. |
| Sex | Male | Male | Male | .. |
| Epidemiological contact history | Yes | Yes | Yes | 3(100%) |
| Chronic medical illness | Prostatic hyperplasia | None | Chronic bronchitis | 2(66.7%) |
| **Symptoms and signs** |  |  |  |  |
| Fever | Yes | Yes | Yes | 3(100%) |
| Dyspnoea | Yes | Yes | Yes | 3(100%) |
| Cough | No | Yes | No | 1(33.3%) |
| Myalgia or fatigue | No | Yes | No | 1(33.3%) |
| Diarrhoea | Yes | No | No | 1(33.3%) |
| Chest pain | No | No | No | 0 |
| Headache | No | No | No | 0 |
| Rhinorrhoea | No | No | No | 0 |
| Sore throat | No | No | No | 0 |
| **Inflammation** |  |  |  |  |
| C-reactive protein (mg/L; normal range 0.0–10.0) | 40↑ | 86.7↑ | 60.7↑ | 3(100%) |
| Procalcitonin (ng/mL; normal range <0.1) | 0.087 | 1.69↑ | 0.115↑ | 2(66.7%) |
| White blood cell count ($\times 10^9$ cells per L; normal range 3.5–9.5) | 7.94 | 4.69 | 5.17 | 0 |
| Neutrophil count ($\times 10^9$ cells per L; normal range 1.8–6.3) | 6.13 | 3.75 | 4.3 | 0 |
| Lymphocyte count ($\times 10^9$ cells per L; normal range 1.1–3.2) | 1.4 | 0.55↓ | 0.6↓ | 2(66.7%) |
| Monocyte count ($\times 10^9$ cells per L; normal range 0.1–0.6) | 0.4 | 0.39 | 0.25 | 0 |
| **Organs function** |  |  |  |  |
| Hypersensitive troponin I (ng/mL; normal range 0.0–0.04) | 0.012 | 0.06↑ | 0.011 | 1(33.3%) |
| Alanine aminotransferase (U/L; normal range 9.0–50.0) | 36 | 19 | 102↑ | 1(33.3%) |
| Aspartate aminotransferase (U/L; normal range 15.0–40.0) | 33 | 60↑ | 122↑ | 2(66.7%) |
| Total bilirubin (μmol/L; normal range 0.0–23.0) | 13.7 | 12.1 | 7 | 0 |
| Urea (mmol/L; normal range 3.1–8.0) | 5.1 | 14.5↑ | 3.7 | 1(33.3%) |
| Creatinine (μmol/L; normal range 57.0–97.0) | 73 | 163↑ | 52↓ | 2(66.7%) |
| **Treatment** |  |  |  |  |
| Antiviral treatment | Yes | Yes | Yes | 3(100%) |
| Antibiotic treatment | Yes | Yes | Yes | 3(100%) |
| Antifungal treatment | No | No | No | 0 |
| Intravenous immunoglobulin therapy | Yes | No | No | 1(33.3%) |
| **Clinical outcome** |  |  |  |  |
| Hospitalisation | No | No | No | 0 |
| Discharge | No | No | No | 0 |
| Died | Yes | Yes | Yes | 3(100%) |
| **Gene loci of 2019-nCoV** |  |  |  |  |
| 2019n-CoV open reading frame 1ab (2019n-CoV ORF1ab) | positive | positive | positive | 3(100%) |
| 2019n-CoV nucleocapsid protein gene (2019n-CoV N gene) | positive | positive | positive | 3(100%) |

### Dynamic changes of immune cells and oximetry saturation surveillance

To determine the dynamic changes of different immune cells in all patients, we analyzed the levels of CD3+ T cells, CD4+ T cells, CD8+ T cells, CD19+ B cells and CD16+56+ NK cells in the peripheral blood of three patients at different time points during the hospitalization. As shown in Figure 1A, Patient 1 had sustained decrease in all types of immune cells during the course. Specifically, CD3+, CD4, CD8+ T cells counts were always below normal range, and CD19+ B cells count dropped below normal range from day 12 and CD16+56+ NK cells count dropped below normal range from day 16. Simultaneously, Patient 1 received oxygen support through oxygen mask (5 L/min) and even high flow oxygen (Figure 1A) due to hypoxemia. However, oximetry saturation (SaO_2_) were still below 95% and not recovered. On day 15, invasive mechanical ventilation was used on him due to sudden respiratory failure, while it didn’t work and the patient died at day 24. Patient 2 had sustained decrease in all types of immune cells at the first six days below normal range (Figure 1B) and hypoxemia. Oxygen mask and high flow oxygen were used to correct the hypoxemia, but the SaO_2_ was almost decreased along the way from day 6 (Figure 1B). Eventually, Patient 2 died at day 13. In Patient 3, CD3+, CD4+, CD8+ T cells were obviously decreased below normal range and CD3+, CD4+ T cells presented a sustained decline from day 4 (Figure 1C). Additionally, during the oxygen support (oxygen mask, > 5 L/min), the SaO_2_ of this patient was oddly declined. From day 8, noninvasive mechanical ventilation was used but didn’t work (Figure 1C). The patient presented a progressive and refractory hypoxemia, and died on day 12. From these results, we found that all patients had immunodepletion and refractory hypoxemia. Interestingly, the phenomenon of immunodepletion appeared earlier than refractory hypoxemia in patients during the progression of COVID-2019 pneumonia.

**Figure 1.**
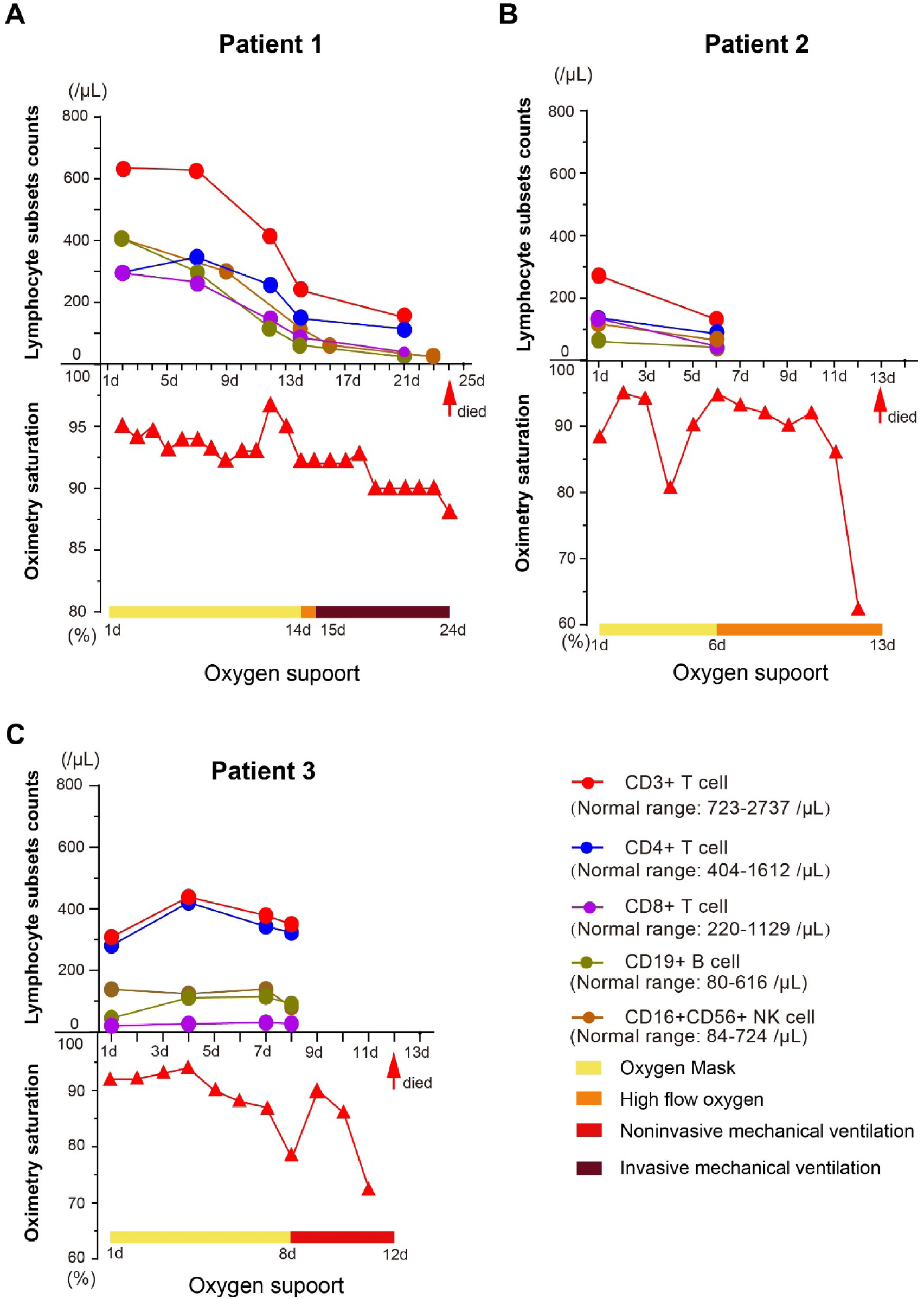
Kinetic analysis of different immune cells counts and oximetry saturation of three patients. (A) Dynamic changes of different immune cells and oximetry saturation in Patient 1; (B) Dynamic changes of different immune cells and oximetry saturation in Patient 2; (C) Dynamic changes of different immune cells and oximetry saturation in Patient 3. Red arrow: death time. Horizontal ordinate: hospital day.

**Figure 2.**
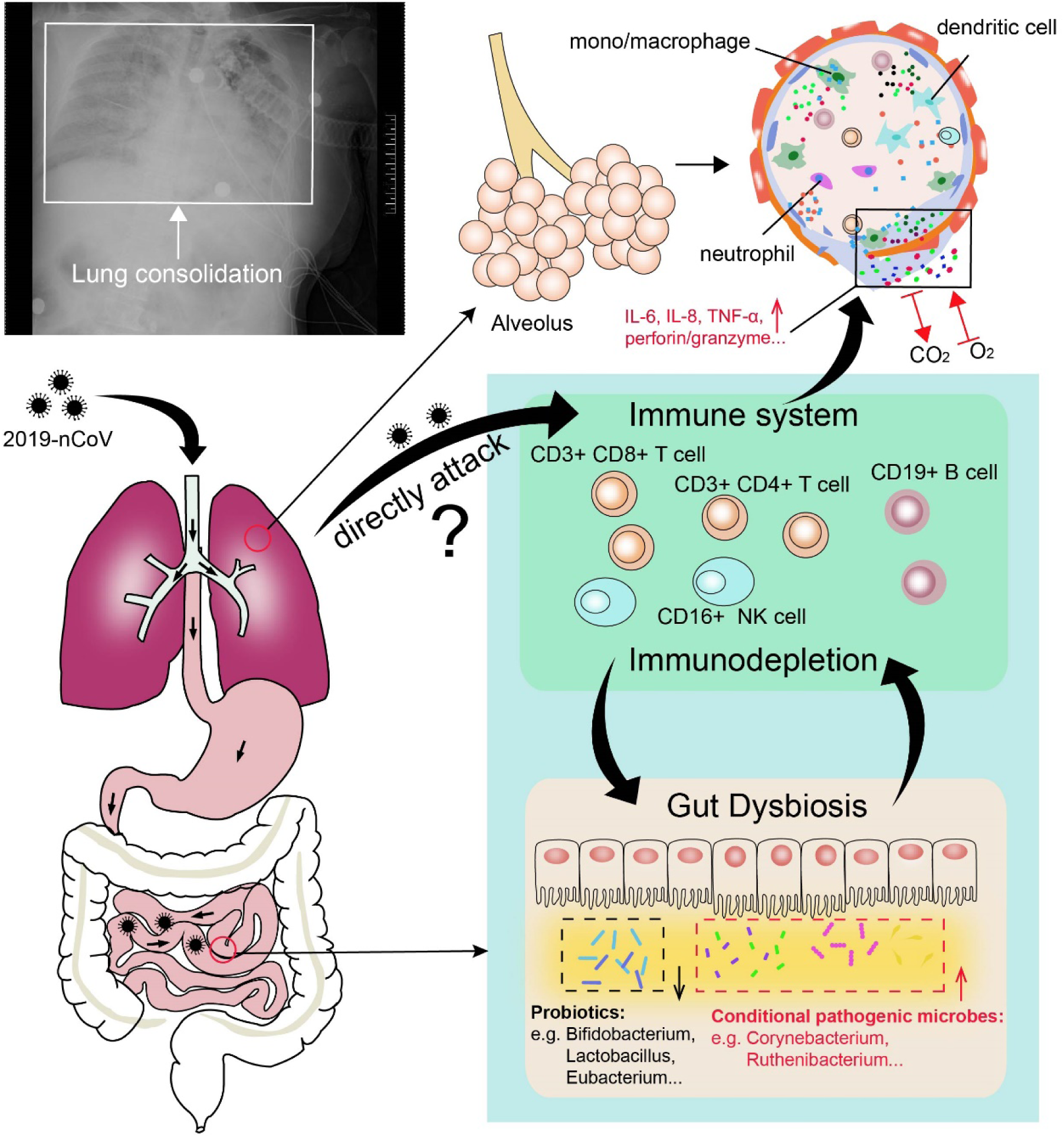
Illustration of potential pathogenic mechanisms of 2019-nCoV.

### Analysis of gut microbes of COVID-19 patients

Through the NTS test, we analyzed the gut microbiota at the level of phylum and genus on 3 anal swab samples from Patient 1 and Patient 3. The data of healthy human intestinal microbiota were imported from Arumugam M’s research^5^. The analysis results showed that no typical pathogens detected in the two patients, such as Clostridium difficile, however the structure of the gut microbiota fluctuated greatly both in bacteria and fungi (Table 2). Firstly, Patient 1’s Actinobacteria was significantly higher than health cohort both in the test of two days in a row. The proportion of Actinobacteria in bacteria of patient 1 for two consecutive days are 70.7% and 44.17%, respectively. The abundance of Actinobacteria increase in patient 1 was mainly triggered by the significant increase in Corynebacterium (77.12%, 44.67%) at the level of genus. In addition, Kluyveromyces dominates the intestinal fungal flora of this patient, which the proportion of fungi for 2 day are 93.4% and 99.26%. Similarly, the proportion of Firmicutes in bacterial microbiota of patient 3 is as high as 81.72% mainly because of the increase of Ruthenibacterium (35.12%) at the genus level, which is significantly higher than that of health cohort (P<0.0001). At the same time, the Aspergillus (59.41%) dominates the intestinal fungal flora of this patient. Overall, all data are significantly different from healthy human gut microbiota (P <0.001).

**Table 2.**
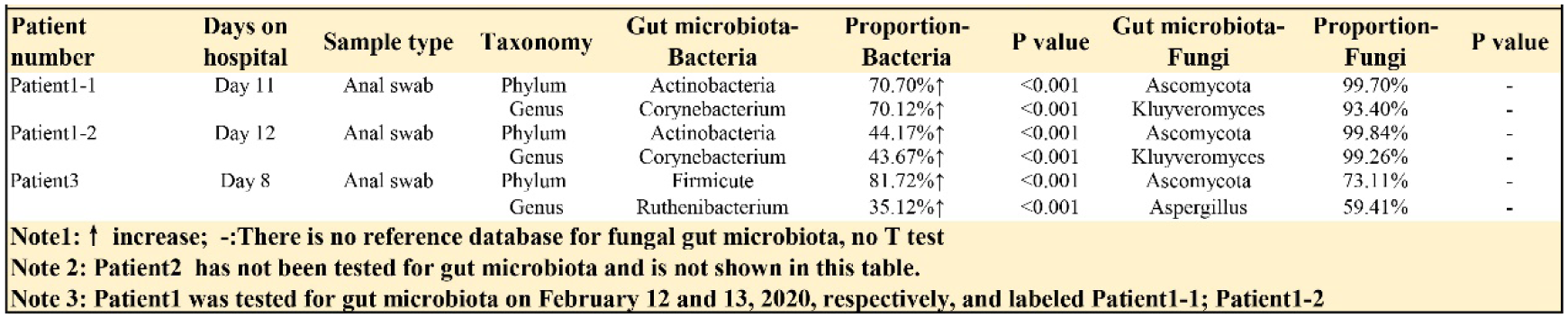
Analysis of gut microbiota in 2 critical patients infected with 2019-nCoV.

## Discussion

In this study, we revealed the clinical characteristics of 3 critical patients with COVID-2019, and found that (1) hypoxemia severity was strongly correlated to the expression levels of immune cells, which indicated that immuno-depletion may well be a key risk factor in the development of COVID-2019, and (2) the imbalance of gut microbiota was also found in these critical patients, which may lead to microbial translocation and a second infection.

The balance between virus and host immunity is closely associated with the outcome of an infection. Clinical studies have shown that coronavirus infection severity was linked to patient health status and an immunocompromised state carried an increased risk of death^6, 7^. It was also demonstrated that the immunosuppressed rhesus macaques had markedly increased MERS-Coronavirus replication in the lung, suggesting the inability in virus clearance^8^. Although the phenomenon of decreased lymphocytes in the periphery blood of COVID-2019 patients has been described in the latest 7th edition of the Diagnosis and Treatment of New Coronavirus Pneumonia published by the National Health Commission of China^9^, the dynamic changes of immune cells and the relation between levels of immune cells and the severity of hypoxemia are still unknown. Consistently, in these critical patients with COVID-2019, we found that the lower immune cell levels, the more serious hypoxia, which suggested that the imbalance between host immunity and coronavirus would lead to worse clinical outcome. Furthermore, evidence have identified the adverse effects of coronavirus on host immune system^10, 11^. Thus, reasons of the immuno-depletion state in these critical patients may be associated with excessive consumption of immune cells, as well as virus-induced immune organs injury.

Multiple studies have demonstrated the interaction between immune system and gut microbiota, and gut microbiota homeostasis is modulated by the immune system^12^. It was shown that a variety of immune factors was capable of altering gut microbiota homeostasis balance, such as IL-10, IL-23, NLRP6, NOD-like receptor protein 3, and IgA^13^. And the imbalance or dysbiosis in gut microbiota would contribute to an enhanced mucosal permeability and thus leading to microbial translocation and a second infection. Moreover, gut microbiota was able to modulate the immune response and played an important role in immune-mediated diseases, suggesting a vicious circle between immune disorder and gut microbiota imbalance^14^. All these findings indicated that the balance of gut microbiota can be disrupted in case of coronavirus infection due to the immune disorder. In this study, we further identified the changes in gut microbiota during the novel coronavirus infection and found that (1) the proportion of probiotics was significantly reduced, such as Bifidobacterium, Lactobacillus, and Eubacterium, and (2) the proportion of conditioned pathogenic bacteria was significantly increased, such as Corynebacterium of Actinobacteria and Ruthenibacterium of Firmicutes. In addition, we also found that the fungi in the gut microbiota of the two patients are not common fungi in healthy cohort (eg. Candida and Saccharomyces cerevisiae), some rare fungi dominate the fungal of gut microbiota, such as Aspergillus, Kluyveromyces. Insight into which species of the microbiota are altered by immune-depletion associated with novel coronavirus infection can help us have a better understanding of the characteristics of COVID-2019.

Based on our findings, we suggested that the early interventions of host immune system and gut microbiota are necessary for COVID-2019 patients with refractory hypoxia. For these critical patients, extracorporeal membrane oxygenation (ECMO) therapy also should be supported early to ameliorate the symptom of refractory hypoxia. In conclusion, we discussed the dynamic characteristics of host immune system and the imbalance of gut microbiota in 3 critical patients with COVID-2019. Hypoxemia severity was closely related with host immune cell levels, and the vicious circle between immune disorder and gut microbiota imbalance may be a high risk of fatal pneumonia.

## Data Availability

Please contact the corresponding authors by e-mail for data availability and permission before reusing any data.

## Acknowledgement

This work was supported by grants from the National Key R&D Program of China [2017YFC1307800].

